# High-resolution mapping of *DMD* duplications using long-read sequencing enables precise carrier screening for Duchenne muscular dystrophy

**DOI:** 10.1101/2025.08.11.25333458

**Authors:** Jing Yang, Yanling Dong, Zhigang Wang, Xinyao Sun, Nana Song, Shanshan Gu, Xue Zhang, Yaya Guo, Xiangzhong Sun, Shiping Chen, Jian Wang, Jiale Xiang

## Abstract

**Purpose:** Exon-level duplications in the *DMD* gene present interpretive challenges due to limitations in resolving their genomic context. We aimed to assess the utility of long-read genome sequencing (lrGS) in characterizing *DMD* duplications and guiding clinical interpretation.

**Methods:** We applied low coverage lrGS (3–10× depth; ∼8.2 kb mean read length) to 18 individuals with *DMD* duplications identified via short-read sequencing. Structural variant calling and breakpoint localization were validated by Sanger sequencing. In addition, the genomic characteristics of the duplication breakpoints were systematically analyzed.

**Results:** lrGS resolved duplication architecture in all cases. Two duplications (11%, 2/18) were extragenic and reclassified as benign; 16 (89%, 16/18) were tandem events within *DMD*. Among tandem duplications, 50% (8/16) were classified as pathogenic/likely pathogenic and 50% (8/16) as variants of uncertain significance. Breakpoints were consistently located in intronic regions, often flanked by repetitive elements.

**Conclusion:** Low-coverage lrGS enables high-resolution mapping of *DMD* duplications and improves variant classification. This approach addresses a key gap in carrier screening and molecular diagnosis of dystrophinopathies, and provides lrGS as a prototype for decoding duplication architecture of monogenic disorders, which is a critical advance in genetic diagnosis.

## Introduction

The *dystrophin* (*DMD*) gene, the largest gene documented in the human genome, encodes dystrophin, a cytoskeletal protein essential for maintaining the structural integrity and functional stability of muscle fibers.^1,2^ Variations in the *DMD* gene cause X-linked recessive neuromuscular disorders including Duchenne muscular dystrophy (DMD), Becker muscular dystrophy (BMD), and X-linked dilated cardiomyopathy, which threaten human health significantly.^3,4^ DMD exhibits a high global incidence, with reported rates exceeding 15 cases per 100,000 live male births in the USA and the UK.^5^ Due to the X-linked recessive inheritance pattern, female carriers have a 50% chance of transmitting the mutation to their sons, while female carriers themselves may also experience milder symptoms with a low probability.^6^ Thus, *DMD* carrier screening is pivotal for reducing DMD incidence.

There are multiple variant types in the *DMD* gene, such as exon deletions (∼68%), duplications of the *DMD* gene (10%), and small mutations (22%).^7^ Many methods, including multiplex ligation-dependent probe amplification (MLPA), quantitative polymerase chain reaction (qPCR), exome sequencing and other techniques, had been developed to detect *DMD*.^8^ None of them can accurately detect duplications (DUP), though the genomic location of duplicated sequence is of importance for its pathogenicity.^9^ With the development of long-read sequencing technologies, such as CycloneSEQ,^10^ Oxford Nanopore Technologies (ONT),^11^ and Pacific Biosciences single-molecule real-time sequencing,^12^ the genomic location of duplicated sequence came into reality. However, with the increase in sequencing depth, the cost of long-read sequencing also rises, and its sequencing accuracy still needs improvement.^13^ Therefore, exploring how to achieve low-cost and high-accuracy diagnosis of *DMD* structural variants has important clinical value.

Here, we developed a detailed protocol integrating short- and long-read genome sequencing for precise analysis of *DMD* duplications (Supplementary Figure 1). Using this approach, we achieved 100% breakpoint detection across 18 cases at sequencing depths ranging from 3× to 10×. Our results demonstrate that this protocol enables accurate breakpoint mapping and facilitates precise variant classification, addressing a critical gap in the interpretation of DMD duplications.

## MATERIALS AND METHODS

### Study design and ethnic statement

Female carriers with *DMD* duplications were identified through expanded carrier screening test using short-read sequencing at the clinical laboratory of Beijing Genomic Institution, China. Duplications were confirmed by MLPA or qPCR (Supplementary Table 1). Written informed consent was obtained from all participants for anonymized scientific use. This study was approved by the institutional review board of the ethnic committee (IRB25049).

**Table 1.** The precise interpretation of *DMD* duplication by short and long read genome sequencing

| Case ID | Dup. region | Length (bp) | Exon | CDS (bp) | Breakpoint location |  | Type | Disrupted ORF | ACMG/AMP criteria | Pathogenicity |
| --- | --- | --- | --- | --- | --- | --- | --- | --- | --- | --- |
| 1 | Retrocopies<br>EX29-37; 39-40 | 1914 | EX29-37;<br>39-40 | 1664 | Chr13:7134<br>6617 | Intergenic region | Non-tandem | No | / | B |
| 2 | ChrX:31329852<br>-31581111 | 251421 | EX56-61 | 946 | ChrX:35927<br>062 | *CFAP47* | Non-tandem | No | / | B |
| 3 | ChrX:32500895<br>-33131827 | 630933 | EX2-19 | 2349 | ChrX:33131<br>827 | *DMD*<br>intron | Tandem | No | PM4+PM2 | VUS |
| 4 | ChrX:32363397<br>-32381229 | 17833 | EX34-36 | 480 | ChrX:32381<br>229 | *DMD*<br>intron | Tandem | No | PM2 | VUS |
| 5 | ChrX:32467042<br>-32654145 | 187104 | EX10-23 | 2202 | ChrX:32654<br>145 | *DMD*<br>intron | Tandem | No | PM2 | VUS |
| 6 | ChrX:30951356<br>-31884895 | 933540 | EX48-79 | 4146 | ChrX:31884<br>895 | *DMD*<br>intron | Tandem | No | PM2 | VUS |
| 7 | ChrX:31384548<br>-31676000 | 291453 | EX54-60 | 1212 | ChrX:31676<br>000 | *DMD*<br>intron | Tandem | No | PM2 | VUS |
| 8 | ChrX:31384548<br>-31676000 | 291453 | EX54-60 | 1212 | ChrX:31676<br>000 | *DMD*<br>intron | Tandem | No | PM2 | VUS |
| 9 | chrX:32559305-32818238 | 258934 | EX6-16 | 1635 | ChrX:32818<br>238 | *DMD*<br>intron | Tandem | No | PM4+PM2 | VUS |
| 10 | chrX:30946034-31153469 | 207436 | EX75-79 | 505 | ChrX:31153<br>469 | *DMD*<br>intron | Tandem | No | PM2 | VUS |
| 11 | ChrX:32070688<br>-32363027 | 292340 | EX37-44 | 1284 | ChrX:32363<br>027 | *DMD*<br>intron | Tandem | No | PS4+PM2+P<br>P4 | LP |
| 12 | ChrX:32044724<br>-32479390 | 434667 | EX22-44 | 3635 | ChrX:32479<br>390 | *DMD*<br>intron | Tandem | Yes | PVS1_Strong<br>+PS4+PM2 | P |
| 13 | ChrX:32108164<br>-32781797 | 673634 | EX8-44 | 5789 | ChrX:32781<br>797 | *DMD*<br>intron | Tandem | Yes | PVS1_Strong<br>+PS4+PM2 | P |
| 14 | ChrX:32108164<br>-32781797 | 673634 | EX8-44 | 5789 | ChrX:32781<br>797 | *DMD*<br>intron | Tandem | Yes | PVS1_Strong<br>+PS4+PM2 | P |
| 15 | ChrX:31254748<br>-31542864 | 288117 | EX56-63 | 1069 | ChrX:31542<br>864 | *DMD*<br>intron | Tandem | Yes | PVS1_Strong<br>+PS4+PM2 | P |
| 16 | ChrX:32044724<br>-32479390 | 434667 | EX22-44 | 3635 | ChrX:32479<br>390 | *DMD*<br>intron | Tandem | Yes | PVS1_Strong<br>+PS4+PM2 | P |
| 17 | ChrX:33005140<br>-33053304 | 48165 | EX2 | 62 | ChrX:33053<br>304 | *DMD*<br>intron | Tandem | Yes | PVS1_Strong<br>+PS4+PM2 | P |
| 18 | ChrX:32380107<br>-32776845 | 396739 | EX8-34 | 4196 | ChrX:32776<br>845 | *DMD*<br>intron | Tandem | Yes | PVS1_Strong<br>+PM2 | LP |

Transcript, NM\_004006.3. Dup, duplication; P, pathogenic; LP, likely pathogenic; VUS, variant of uncertain significance; EX, exon; chr., chromosome; ORF, open reading frame; ACMG/AMP, American College of Medical Genetics and Genomics/Association for Molecular Pathology

### Long-read sequencing and data analysis

Genomic DNA was extracted from peripheral blood using the Magpure Tissue & Blood DNA Kit following the manufacturer’s instructions (Magen, China). DNA integrity was assessed by electrophoresis. LrGS was performed using the CycloneSEQ library preparation kit. Briefly, 2 µg DNA was size-selected with 0.4× VAHTS DNA Clean Beads (Vazyme, China). After end repair, dA-tailing and adaptor ligation of DNA fragments, the library was sequenced on CycloneSEQ platform (MGI, China).^10^

Raw sequencing data were processed by filtering low-quality reads using fastp (v0.23.2) and aligned to the hg38 reference genome with minimap2 (v2.17-r941). For *DMD* duplication analysis, structural variant callers Sniffles2 (v2.4) and cuteSV (v2.1.1) were employed.^14^ Visualization of lrGS data was conducted using the Integrative Genomics Viewer.

### Sanger Sequencing

The breakpoints of the *DMD* gene identified by lrGS were further confirmed via Sanger sequencing. Primer sequences are listed in Supplementary Table 2.

**Table 2.** The characteristics of breakpoint locations of DMD duplications

| Case ID | Breakpoint location | Repetitive elements 1 kb upstream or downstream of the breakpoint |  |  |  |  |  |
| --- | --- | --- | --- | --- | --- | --- | --- |
|  |  | SINE | LINE | LTR | DNA repeat element | Simple repeats | Low complexity repeats |
| 1 | Chr13:71346617 |  |  | * |  | *** | ** |
| 2 | ChrX:35927062 | * | *** |  |  | *** |  |
| 3 | ChrX:33131827 | ** |  |  |  |  |  |
| 4 | ChrX:32381229 | *** |  |  |  | *** |  |
| 5 | ChrX:32654145 |  | * |  |  |  |  |
| 6 | ChrX:31884895 | ** | ** |  |  |  |  |
| 7 | ChrX:31676000 | ** |  |  |  |  |  |
| 8 | ChrX:31676000 | ** |  |  |  |  |  |
| 9 | ChrX:32818238 |  |  | ** |  |  |  |
| 10 | ChrX:31153469 | ** | * |  |  |  |  |
| 11 | ChrX:32363027 |  |  |  |  | ** | ** |
| 12 | ChrX:32479390 |  | * |  |  |  |  |
| 13 | ChrX:32781797 | *** | *** |  | *** |  |  |
| 14 | ChrX:32781797 | *** | *** |  | *** |  |  |
| 15 | ChrX:31542864 | ** | *** | * | *** |  |  |
| 16 | ChrX:32479390 |  | * |  |  |  |  |
| 17 | ChrX:33053304 | * | ** | ** |  | *** |  |
| 18 | ChrX:32776845 |  |  | ** |  |  |  |

SINE, Short Interspersed Nuclear Elements; LINE, Long Interspersed Nuclear Elements. LTR, Long Terminal Repeat. \*□ Indicates that the breakpoint is located within a repetitive element. \*\*□ Indicates that repetitive elements (e.g., SINE, LINE, LTR) are present within 500 bp upstream or downstream of the breakpoint. \*\*\*□ Indicates that repetitive elements are located within 1,000 bp upstream or downstream of the breakpoint but not within 500 bp. Repetitive elements were analyzed by RepeatMasker.

## Results

Across all tested samples, the average read length was 8.2 kb, and sequencing coverage ranged from 3× to 10×. Among 18 individuals previously identified as *DMD* duplication carriers by short-read sequencing, lrGS resolved 2 cases (11%, 2/18) as non-tandem duplications (cases #1 and #2) that were subsequently classified as benign for *DMD* (Table 1). The remaining 16 cases (89%, 16/18) were determined to be tandem duplications (cases #3–18, Table 1). Among these, 50% (8/16) were classified as pathogenic or likely pathogenic, while 50% (8/16) were classified as variants of uncertain significance. Collectively, 55% (10/18) of the duplications revealed by short-read sequencing were ultimately determined to be benign or variant of uncertain significance with respect to DMD.

Of the two non-tandem duplications, the first one (case #1) was a complex retrotransposition event. Short-read sequencing identified a duplication spanning exons 30-40 of the *DMD* gene, excluding exon 38 (Supplementary Table 3). However, MLPA detected the duplication of exon 29 and failed to identify duplications in exons 32, 39, and 40 (Supplementary Table 3). Similarly, qPCR confirmed the duplication of exon 29 but did not detect duplication of exon 31 (Supplementary Tabe 3), indicating discordance among these methods. LrGS identified a retrotransposition event that exons 29–40 (excluding exon 38) were reverse-transcribed from *DMD* mRNA and inserted into chromosome 13 (chr13:71346617, hg38), forming a processed pseudogene that did not disrupt the endogenous *DMD* transcription (Table 1, Supplementary Figure 2). As this retrocopy did not disrupt the *DMD* gene, the variant was classified as benign.

The second non-tandem duplications (case #2) involved exons 56–61 of the *DMD* gene. lrGS revealed that the duplicated sequence (ChrX:31329852-31581111, hg38) was inverted and inserted into intron 2 of the *CFAP47* gene at Xp21.1 (Table 1, Supplementary Figure 3). *CFAP47* is located near *DMD*, with a relatively short genomic distance between the two genes (2.7 Mb apart). PCR and Sanger sequencing confirmed the breakpoint at chrX:35945179 (Table 1, Supplementary Figure 3). Because the insertion occurred outside the *DMD* gene and preserved its reading frame, the variant was classified as benign with respect to DMD.

In addition to above duplications, sixteen tandem duplications were identified, all of which had breakpoints located within *DMD* introns (Table1). Of these, 44% (7/16) were predicted to disrupt the open reading frame (ORF) and likely trigger nonsense-mediated decay (cases #12–18). According to the variant interpretation guidelines proposed by the American College of Medical Genetics and Genomics and the Association for Molecular Pathology,^15,16^ these variants were classified as pathogenic or likely pathogenic. The remaining nine tandem duplications preserved the ORF. Of them, eight were classified as variant of uncertain significance (cases #3–10), and one was classified as likely pathogenic (case #11). Breakpoints of all cases were validated by Sanger sequencing and are illustrated in Supplementary Figures 4–19.

To further investigate the genomic context of *DMD* duplication breakpoints, we analyzed 1,000 bp sequences flanking each junction site (1000 bp upstream or downstream) by RepeatMasker. Notably, repetitive elements, including short interspersed nuclear elements (SINEs), long interspersed nuclear elements (LINEs), and long terminal repeats (LTRs), et al. were frequently observed in these regions (Table 2). In eight cases, the breakpoint was located directly within a repetitive element. In an additional seven cases, repetitive elements were present within 500 bp of the breakpoint. For three cases, repetitive sequences were found within the 1,000 bp window. These findings suggested that repetitive sequences might contribute to the formation of *DMD* duplications by facilitating structural rearrangements through non-allelic homologous recombination or other repeat-mediated mechanisms.

## Discussion

The clinical significance of characterizing exon-level duplications in the *DMD* gene is increasingly recognized. However, a persistent limitation is the inability to detect the genomic architecture of such duplications through conventional assays. In this study, we addressed this gap by developing a lrGS approach (3–10× depth) that leverages extended read lengths (∼8.2 kb) to directly identify duplication breakpoints at nucleotide resolution. This method enabled precise characterization of *DMD* exon-level duplications, establishing lrGS as a prototype strategy for decoding duplication architecture of monogenic disorders and filling a critical void in molecular diagnostics.

Conventional methods—including MLPA, qPCR, and short-read sequencing—fail to determine duplication location or orientation, impeding discrimination between pathogenic and benign events, particularly for ambiguous or extragenic duplications. ^3,17^. In our cohort, 55% of duplications were reclassified as benign or variant of uncertain significance based on lrGS-derived localization, highlighting the limitations of standard assays and the structural complexity of genomic duplications.

Additionally, our findings underscore the necessity of breakpoint localization for accurate pathogenicity assessment of *DMD* duplications (Supplementary Figure 20). Phenotypic outcomes critically depend on whether duplications preserve or disrupt the ORF. Tandem duplications maintaining the ORF typically associate with milder BMD, while frameshifting rearrangements lead to severe DMD phenotypes.^7^ Consequently, breakpoint mapping is indispensable for variant classification and genetic counseling.

Consistent with previous studies^18^, most duplications (89%, 16/18) occur in tandem orientation which preserves canonical splicing patterns and reduces the likelihood of aberrant transcripts^19^. Notably, all tandem duplication breakpoints occurred within intronic regions, being consistent with the genomic structure of *DMD*, which spans >2 Mb with exonic sequences comprising <1% of the locus.^2^ Repetitive elements such as LINEs, SINEs, and other transposable elements were frequently observed near breakpoint junctions, suggesting that non-allelic homologous recombination may underlie duplication formation.^20,21^ These findings are consistent with prior reports and highlight the mechanistic relevance of repetitive elements in structural variant generation.

Extragenic tandem duplications—those occurring outside the *DMD* locus—are generally benign for dystrophinopathy but may confer incidental disease risks if disrupting other genes.^22^ For example, in Case #2, an extragenic duplication inserted into *CFAP47* (a gene linked to ciliopathies) could independently contribute to phenotypic complexity. Conventional methods frequently misclassify such events,^23^ as illustrated by two initially “likely pathogenic” extragenic duplications in our cohort that were benign for DMD.

While lrGS offers advantages over short-read sequencing in structural variant resolution, current limitations include lower base-level accuracy, higher costs, and depth-dependent sensitivity.^24,25^ Our data indicates that a relative low coverage (3-10x) will suffice for tandem duplication breakpoint mapping. This positions low-coverage lrGS as an alternative to patients who were found to carry duplications, especially for those with X-linked disorders, which bring male fetus a 50% risk of suffering the disorder. As lrGS technology evolves, declining costs and improved accuracy may enable broader adoption in genetic disease screening.

Taken together, our study demonstrates that low-coverage lrGS enables accurate breakpoint mapping of exon-level duplications and facilitates precise variant classification. This approach fills a critical gap, particularly in cases where conventional methods yield inconclusive or misleading results. As structural variant interpretation becomes increasingly central to genomic medicine, lrGS offers a scalable and informative option for clinical diagnostics and variant interpretation in neuromuscular and other monogenic disorders.

## Data Availability

The data supporting the conclusions of this article are included within the article and its additional files.

## Author Contributions

Conceptualization: J.X., W.J.; Data Curation: J.Y., X.S., Z.W., Y.D.; Formal Analysis: J.Y., X.S., Z.W.,Y.D.; Investigation: J.Y., N.S., X.S., X.Z.,Y.G.; Methodology: J.X., W.J.; Software: X.S., S.G.; Supervision: J.X., W.J.; Writing–original draft: J.X., Z.W. Writing–review and editing: All authors.

## Ethics Declaration

This study was approved by the institutional review board of the ethnic committee (IRB25049). Written informed consent was obtained from all participants for anonymized scientific use.

