## Supplementary Figures and Tables for "High-resolution mapping of *DMD* duplications using long-read sequencing enables precise carrier screening for Duchenne muscular dystrophy"

**Supplementary Figure 1.** Workflow for precision interpretation of *DMD* duplications assisted by low-depth long-read genome sequencing.

**Supplementary Figure 2-19.** Detection and confirmation of *DMD* gene duplication

**Supplementary Figure 20.** Schematic representations of candidate structural variants resolved using long-read genome sequencing.

**Supplementary Table 1.** Eighteen carriers of *DMD* gene duplications were initially identified by short-read sequencing (SRS) and confirmed by multiplex ligation-dependent probe amplification (MLPA) or qPCR.

**Supplementary Table 2**. Primers used for breakpoints validation

**Supplementary Table 3.** The inconsistency of the MLPA, qPCR, and short-read sequencing test results in Case 1

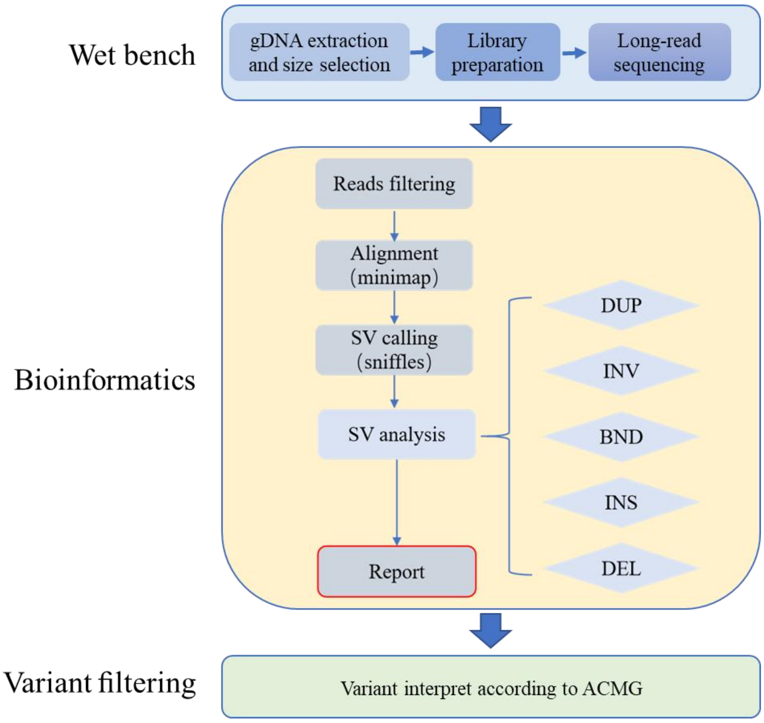


**Supplementary Figure 1.** Workflow for precision interpretation of *DMD* duplications assisted by long-read genome sequencing. SV, Structural Variation; DUP, Duplication; INV, Inversion; BND, Breakend; INS, Insertion; DEL, Deletion.

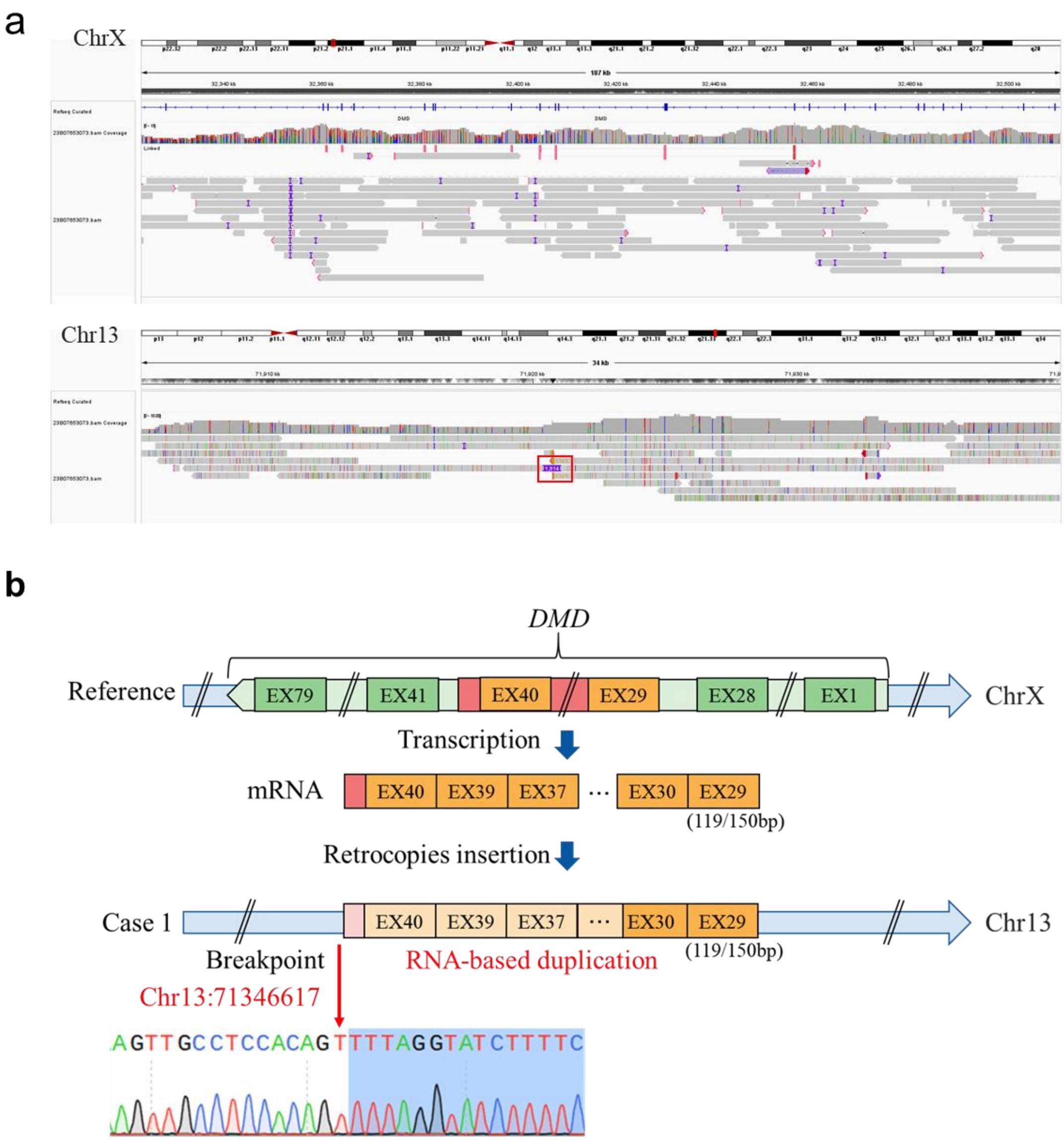


**Supplementary Figure 2.** Detection and confirmation of *DMD* gene duplication in case 1. **A.** Long-read genome sequencing data for case 1 was visualized using the IGV software. There are exon sequences of *DMD* insertion in the intergene region of chromosome 13. The red box points to reads supporting the duplication. **B.** The mRNA of EX29-40 (without EX38) of the *DMD* gene are reverse transcribed to DNA and inserted into the intergenic region of chr13, which was confirmed by Sanger sequencing. The junction of breakpoint was located at Chr13:71346617 (hg38). Chr, chromosome; EX, exon.

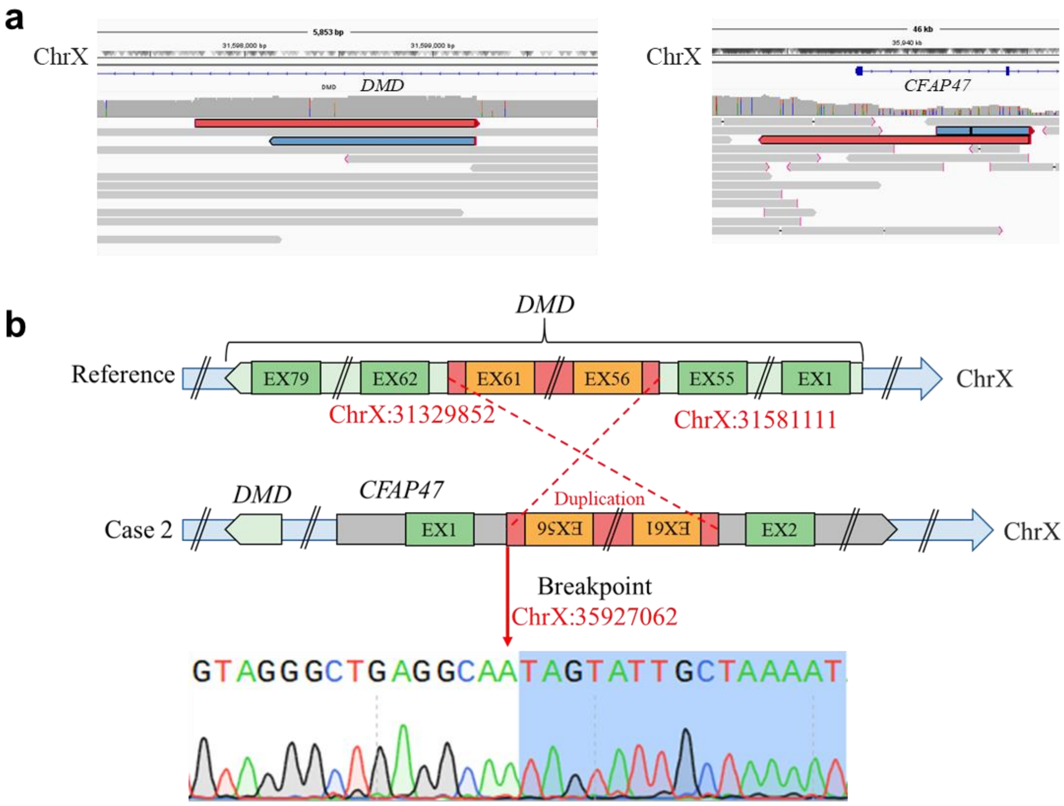


**Supplementary Figure 3.** Detection and confirmation of *DMD* gene duplication in case 2. **A.** Long-read genome sequencing data for case 2 was visualized using the IGV software. Reads supporting the duplication are shown in red or blue. **B.** The inversion duplication of exons 56-61 (ChrX:31329852-31581111, hg38) was inserted between EX1 and EX2 of *CFAP47* at chrX, which was confirmed by Sanger sequencing. The junction of breakpoint was located at chrX:35927062 (hg38). Chr, chromosome; EX, exon.

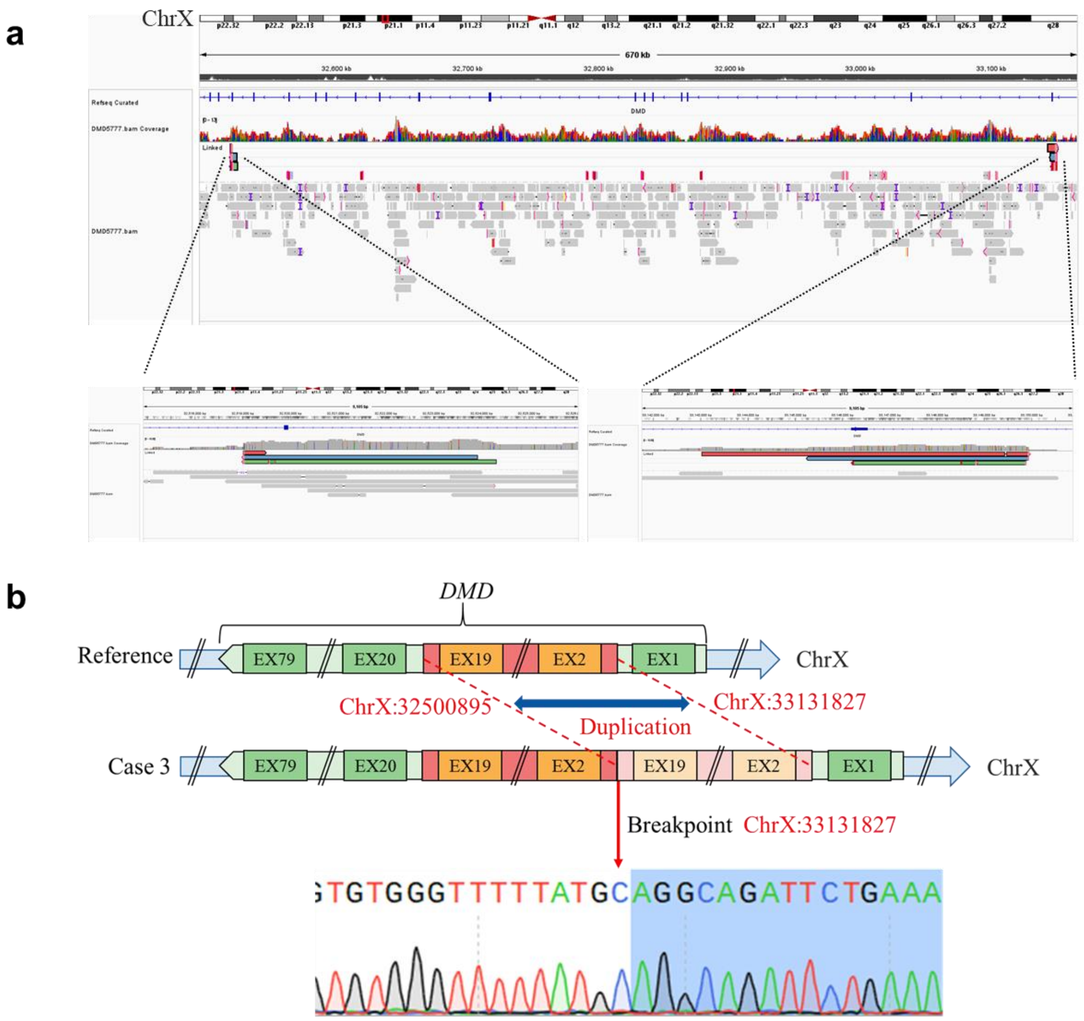


**Supplementary Figure 4**. Detection and confirmation of *DMD* gene duplication in case 3. **A.** Long-read genome sequencing data for case 3 was visualized using the IGV software. Reads supporting the duplication are shown in red, blue or green. **B.** A tandem duplication of exons 2-19 (ChrX:32500895-33131827, hg38) in the *DMD* gene was identified in case 3 by Long-read genome sequencing and Sanger sequencing. The junction of breakpoint was located at chrX:33131827 (hg38). Chr, chromosome; EX, exon.

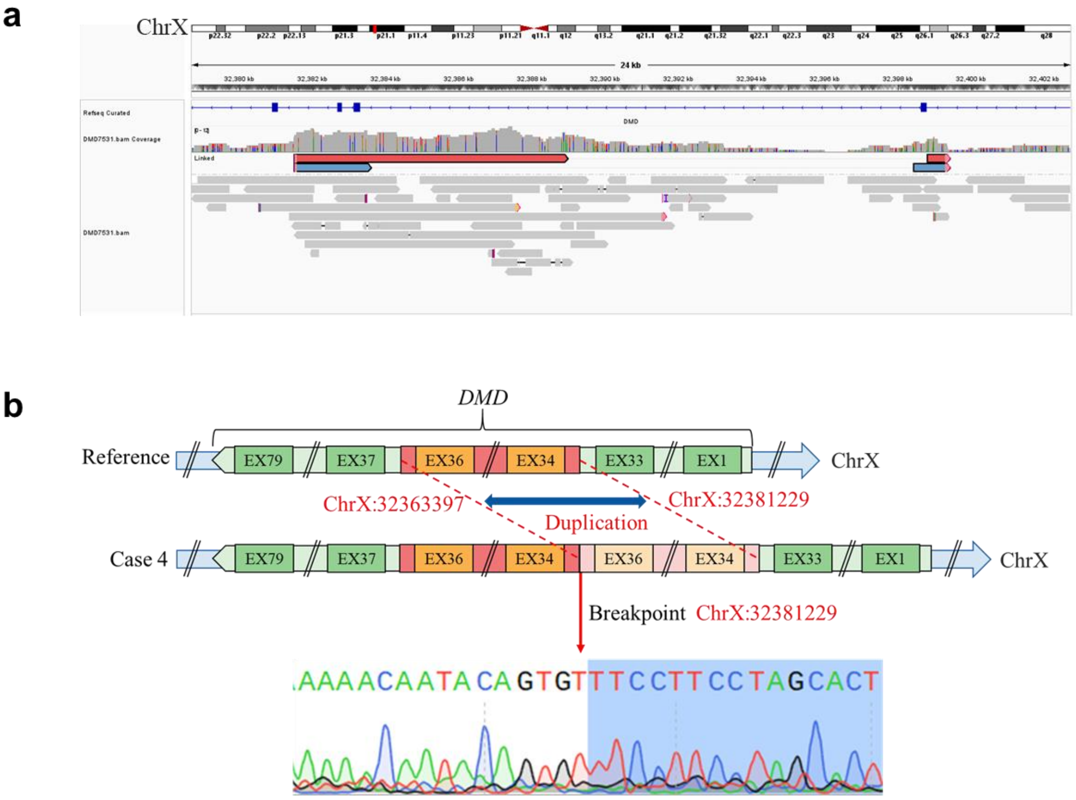


**Supplementary Figure 5**. Detection and confirmation of *DMD* gene duplication in case 4. **A.** Long-read genome sequencing data for case 4 was visualized using the IGV software. Reads supporting the duplication are shown in red or blue. **B.** A tandem duplication of exons 34-36 (ChrX:32363397-32381229, hg38) in the *DMD* gene was identified in case 4 by Long-read genome sequencing and Sanger sequencing. The junction of breakpoint was located at chrX:32381229 (hg38). Chr, chromosome; EX, exon.

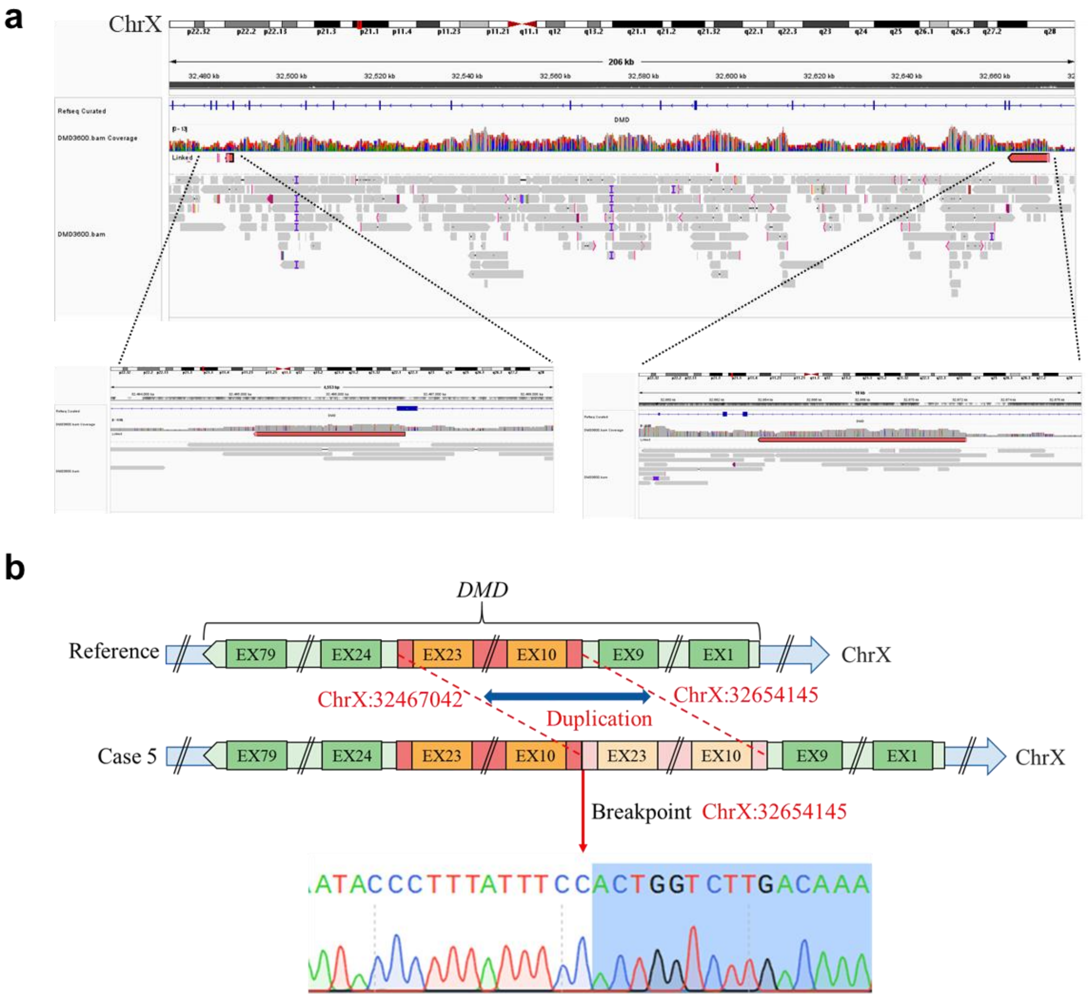


**Supplementary Figure 6**. Detection and confirmation of *DMD* gene duplication in case 5. **A.** Long-read genome sequencing data for case 5 was visualized using the IGV software. Reads supporting the duplication are shown in red. **B.** A tandem duplication of exons 10-23 (ChrX:32467042-32654145, hg38) in the *DMD* gene was identified in case 5 by Long-read genome sequencing and Sanger sequencing. The junction of breakpoint was located at chrX:32654145 (hg38). Chr, chromosome; EX, exon.

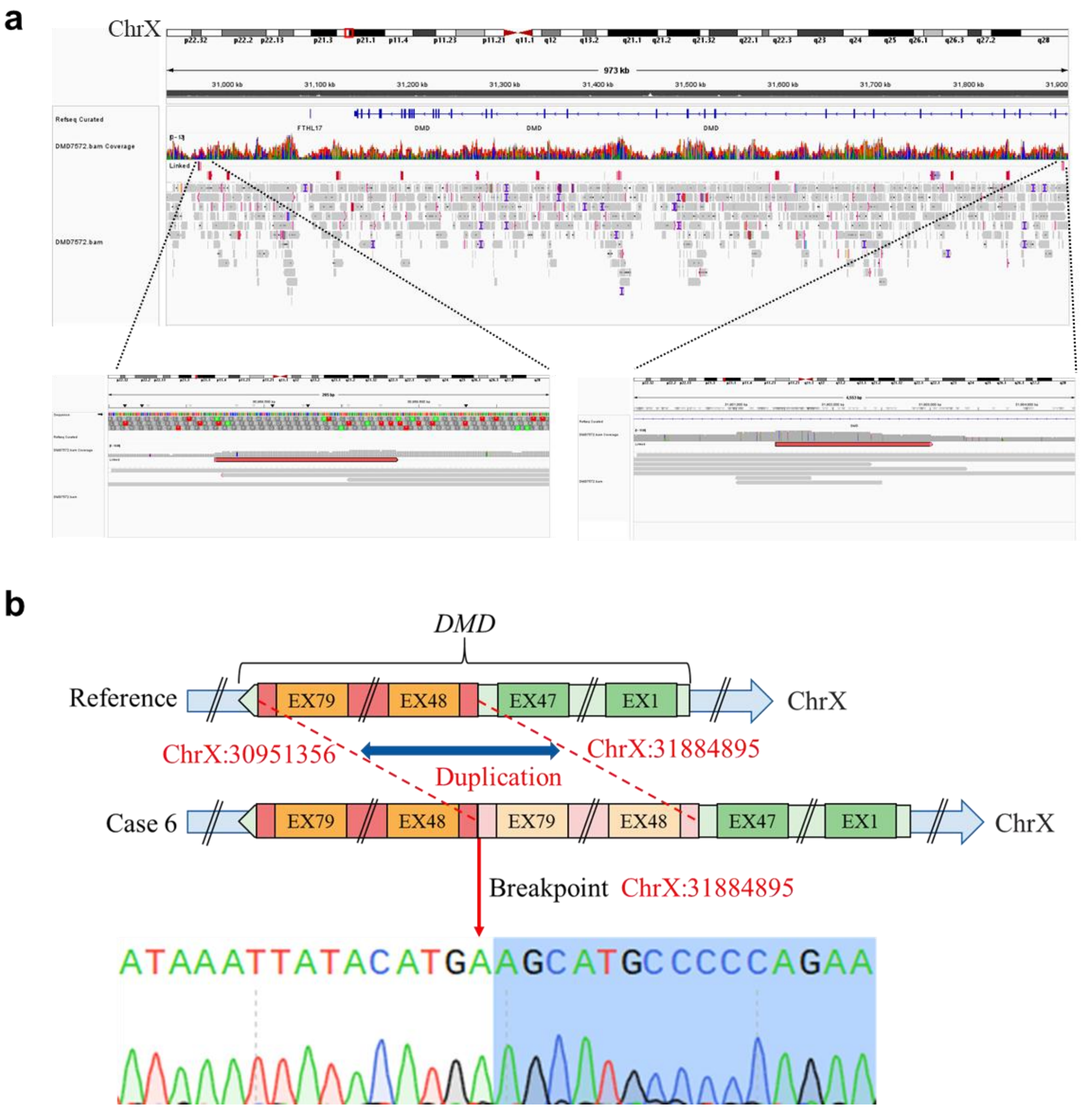


**Supplementary Figure 7**. Detection and confirmation of *DMD* gene duplication in case 6. **A.** Long-read genome sequencing data for case 6 was visualized using the IGV software. Reads supporting the duplication are shown in red. **B.** A tandem duplication of exons 48-79 (ChrX:30951356-31884895, hg38) in the *DMD* gene was identified in case 6 by Long-read genome sequencing and Sanger sequencing. The junction of breakpoint was located at chrX:31884895 (hg38). Chr, chromosome; EX, exon.

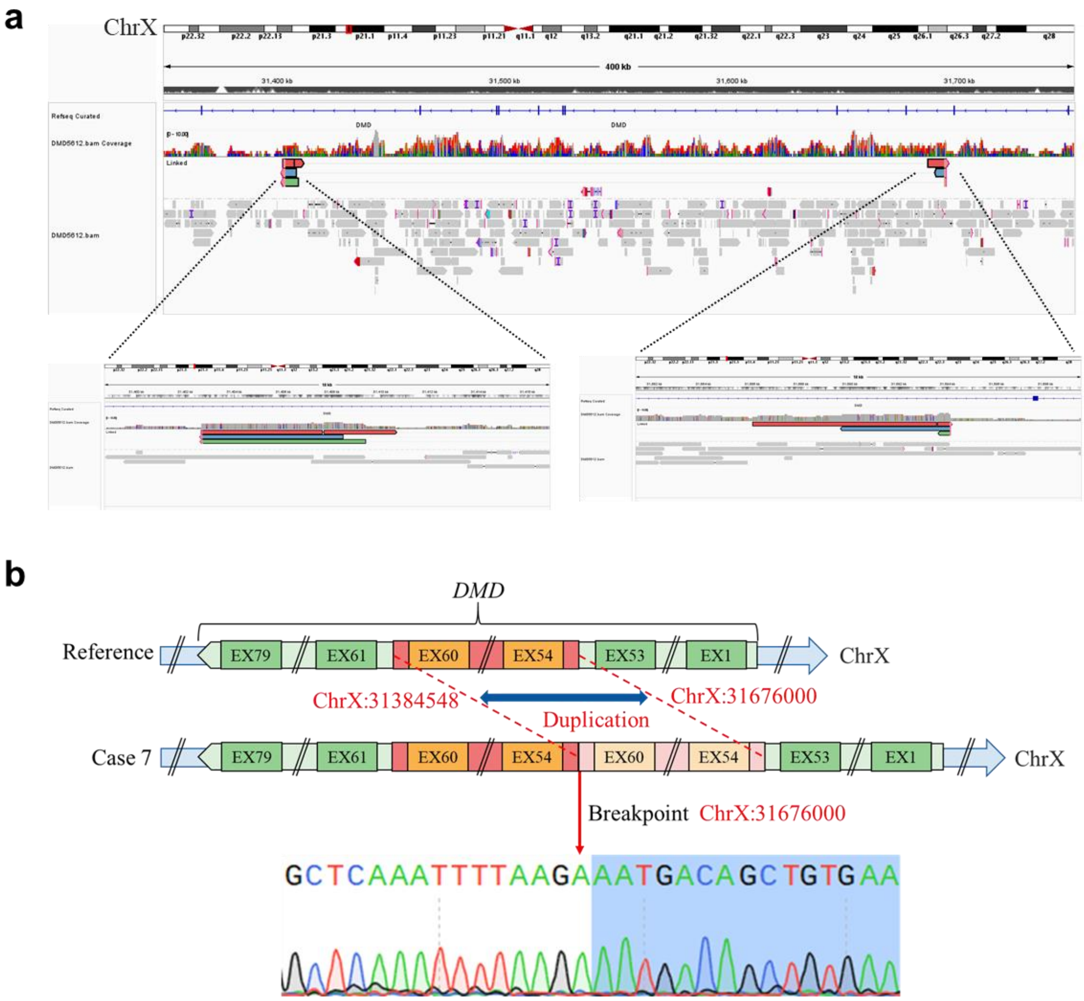


**Supplementary Figure 8**. Detection and confirmation of *DMD* gene duplication in case 7. **A.** Long-read genome sequencing data for case 7 was visualized using the IGV software. Reads supporting the duplication are shown in red, blue or green. **B.** A tandem duplication of exons 54-60 (ChrX:31384548-31676000, hg38) in the *DMD* gene was identified in case 7 by Long-read genome sequencing and Sanger sequencing. The junction of breakpoint was located at chrX:31676000 (hg38). Chr, chromosome; EX, exon.

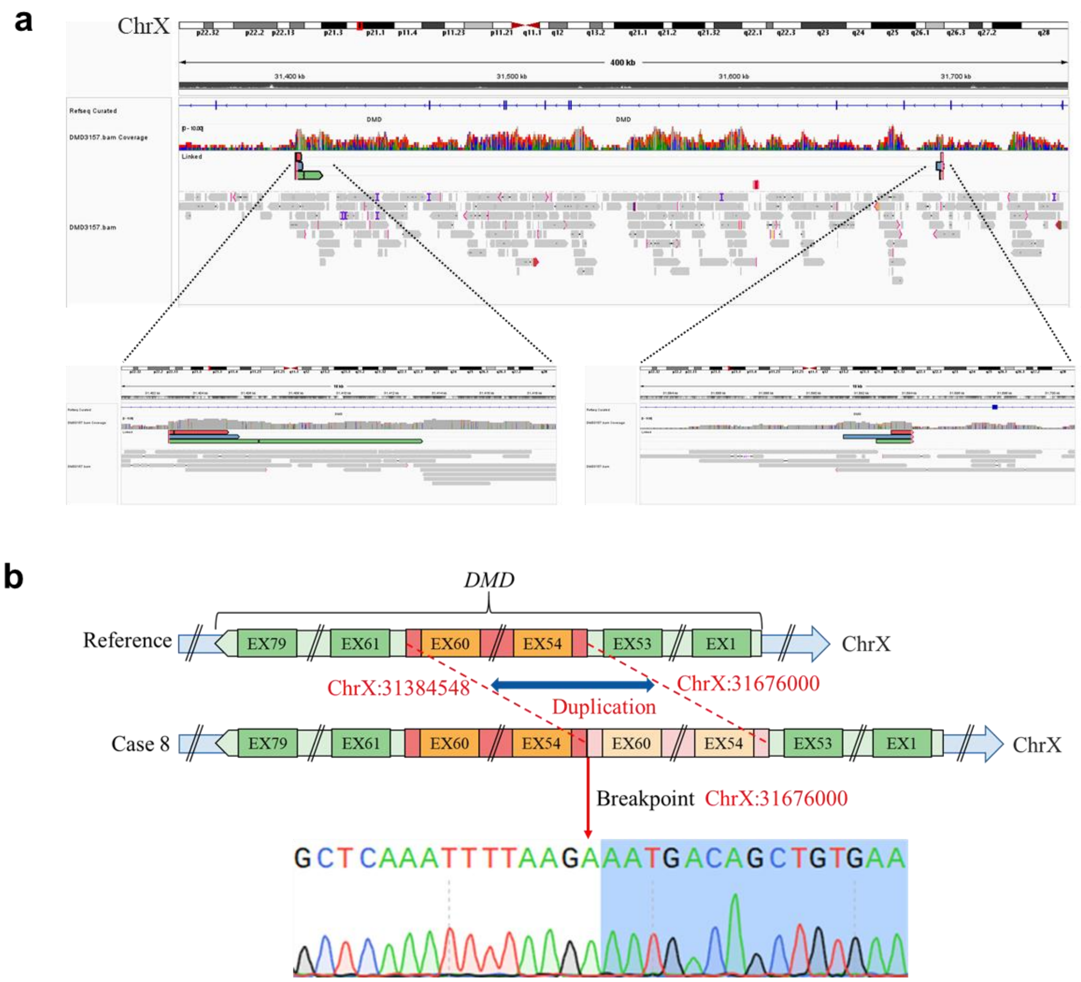


**Supplementary Figure 9**. Detection and confirmation of *DMD* gene duplication in case 8. **A.** Long-read genome sequencing data for case 8 was visualized using the IGV software. Reads supporting the duplication are shown in red, blue or green. **B.** A tandem duplication of exons 54-60 (ChrX:31384548-31676000, hg38) in the *DMD* gene was identified in case 8 by Long-read genome sequencing and Sanger sequencing. The junction of breakpoint was located at chrX:31676000 (hg38). Chr, chromosome; EX, exon.

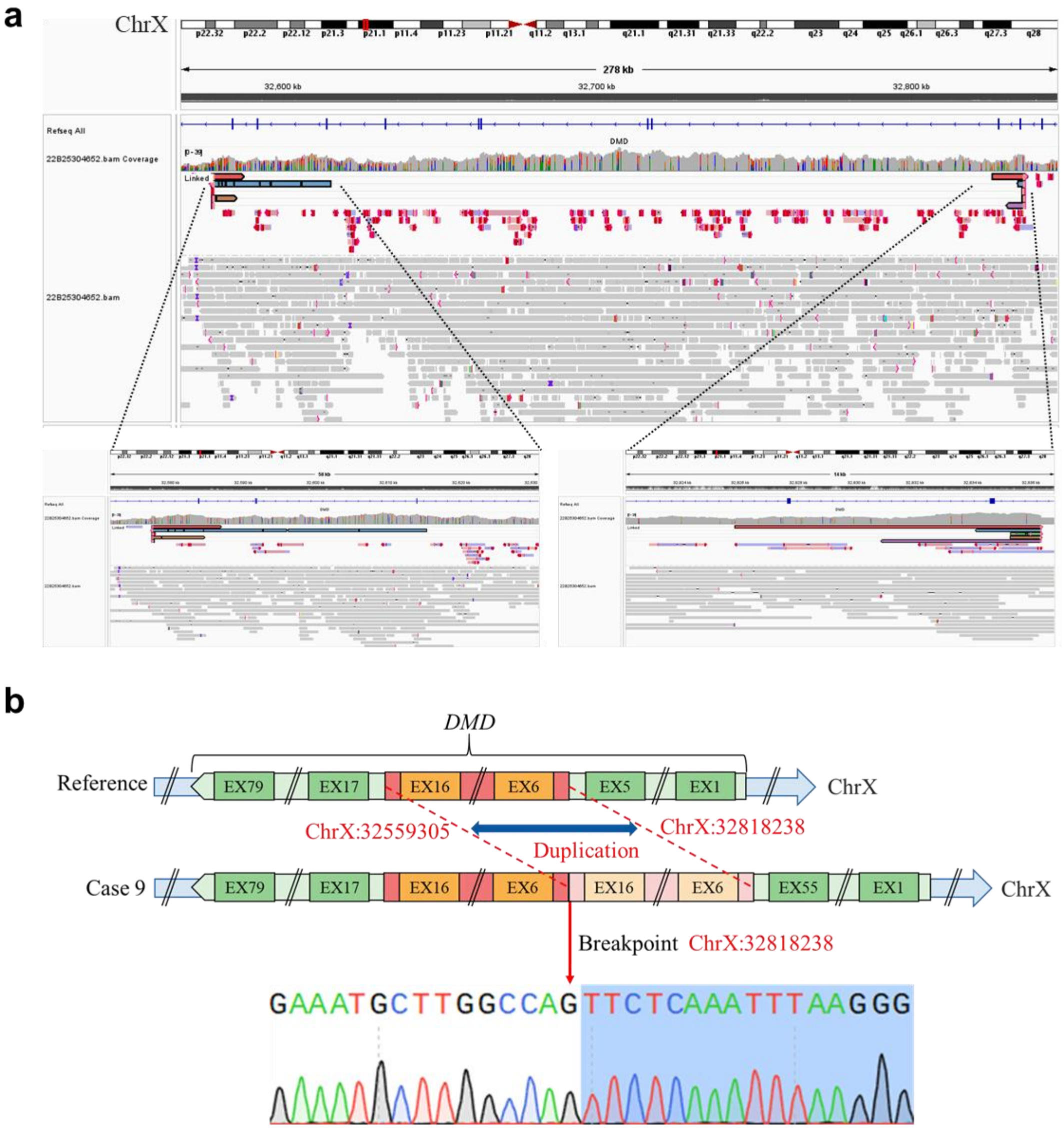


**Supplementary Figure 10**. Detection and confirmation of *DMD* gene duplication in case 9. **A.** Long-read genome sequencing data for case 9 was visualized using the IGV software. Reads supporting the duplication are shown in five different colors. **B.** A tandem duplication of exons 6-16 (chrX:32559305-32818238, hg38) in the *DMD* gene was identified in case 9 by Long-read genome sequencing and Sanger sequencing. The junction of breakpoint was located at chrX: 32818238 (hg38). Chr, chromosome; EX, exon.

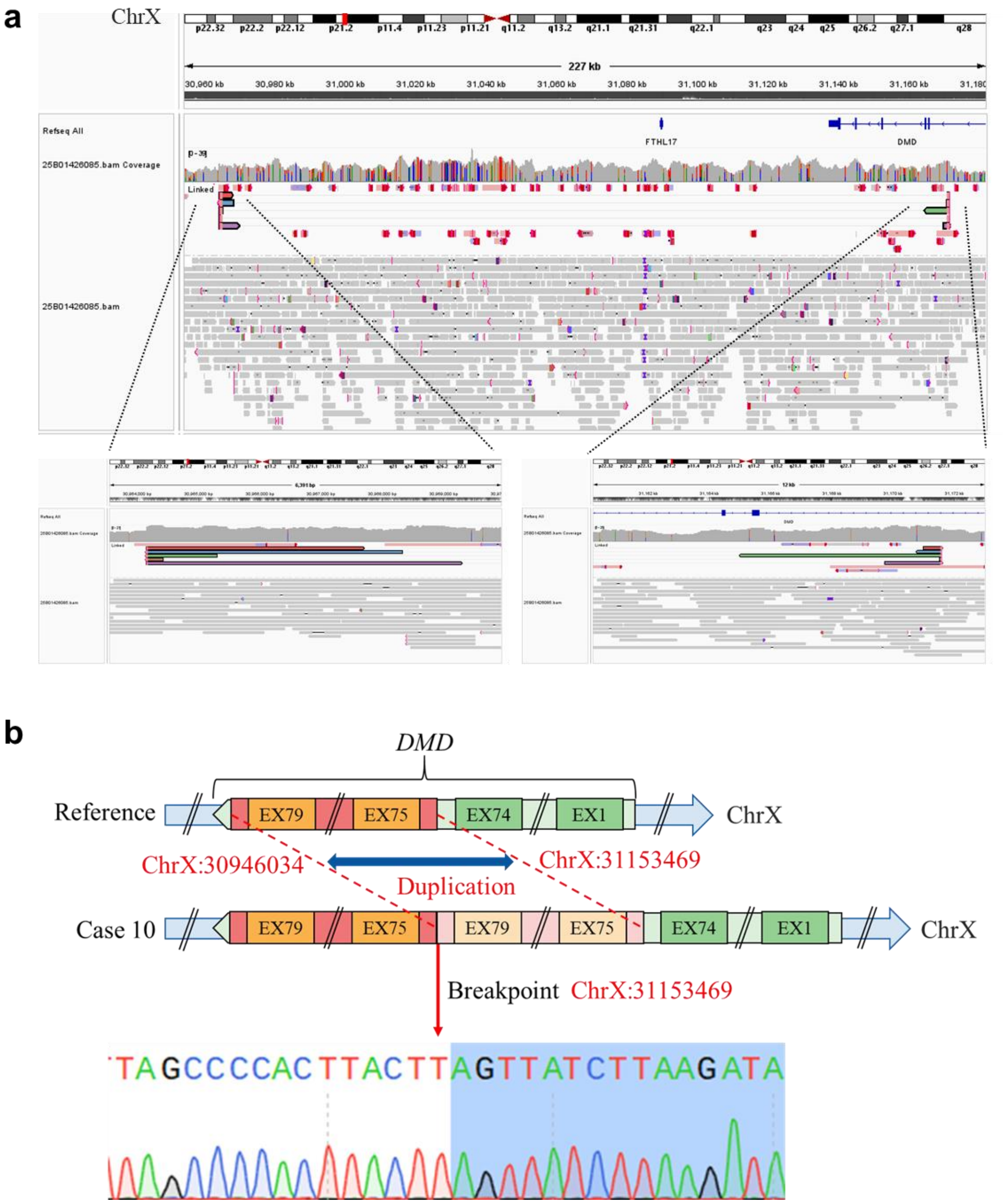


**Supplementary Figure 11**. Detection and confirmation of *DMD* gene duplication in case 10. **A.** Long-read genome sequencing data for case 10 was visualized using the IGV software. Reads supporting the duplication are shown in five different colors. **B.** A tandem duplication of exons 75-79 (chrX:30946034-31153469, hg38) in the *DMD* gene was identified in case 10 by Long-read genome sequencing and Sanger sequencing. The junction of breakpoint was located at chrX: 32818238 (hg38). Chr, chromosome; EX, exon.

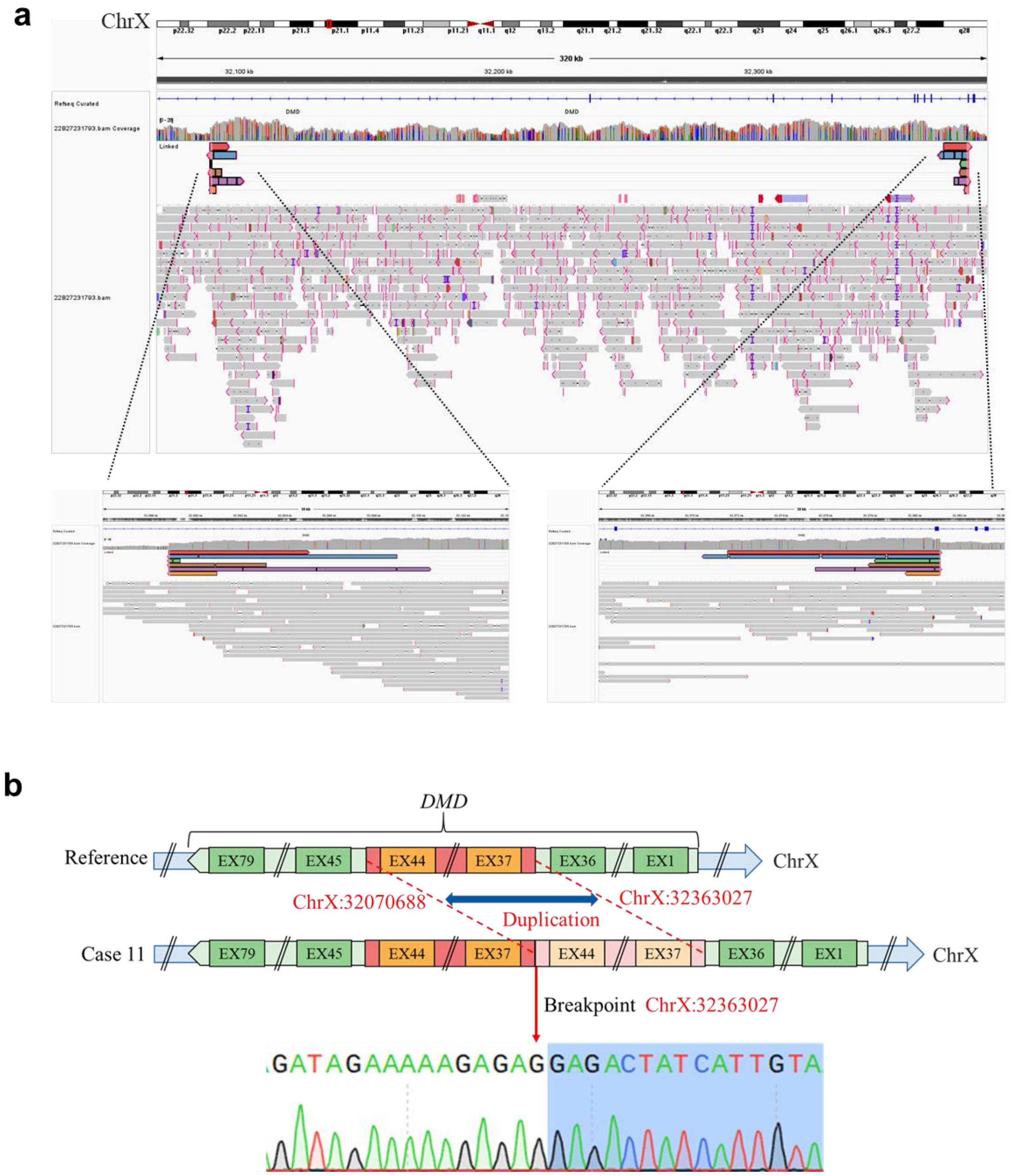


**Supplementary Figure 12**. Detection and confirmation of *DMD* gene duplication in case 11. **A.** Long-read genome sequencing data for case 11 was visualized using the IGV software. Reads supporting the duplication are shown in six different colors. **B.** A tandem duplication of exons 37-44 (ChrX:32070688-32363027, hg38) in the *DMD* gene was identified in case 11 by Long-read genome sequencing and Sanger sequencing. The junction of breakpoint was located at chrX: 32363027 (hg38). Chr, chromosome; EX, exon.

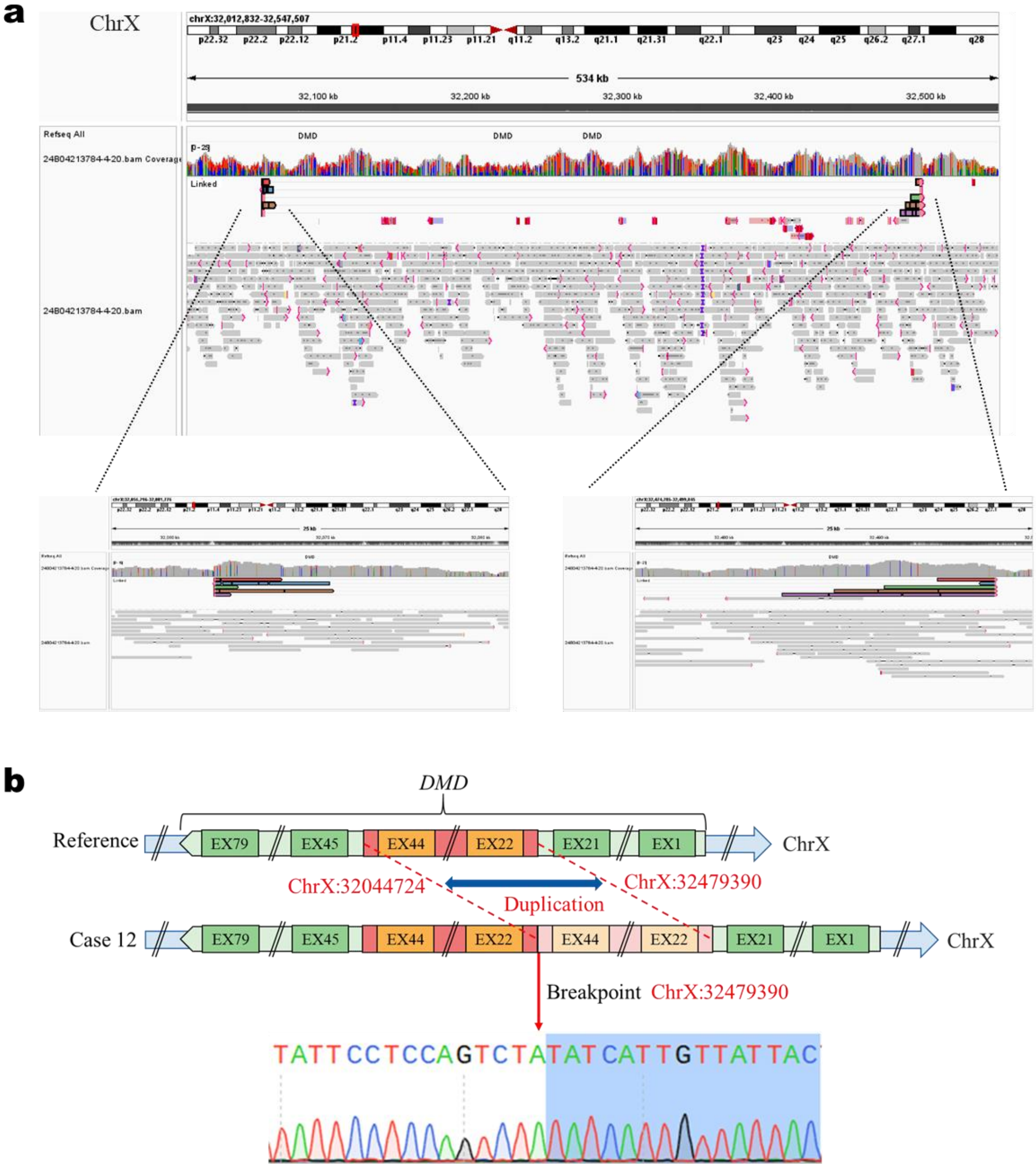


**Supplementary Figure 13**. Detection and confirmation of *DMD* gene duplication in case 12. **A.** Long-read genome sequencing data for case 12 was visualized using the IGV software. Reads supporting the duplication are shown in five different colors . **B.** A tandem duplication of exons 22-44 (ChrX:32044724-32479390, hg38) in the *DMD* gene was identified in case 12 by Long-read genome sequencing and Sanger sequencing. The junction of breakpoint was located at chrX:32479390 (hg38). Chr, chromosome; EX, exon.

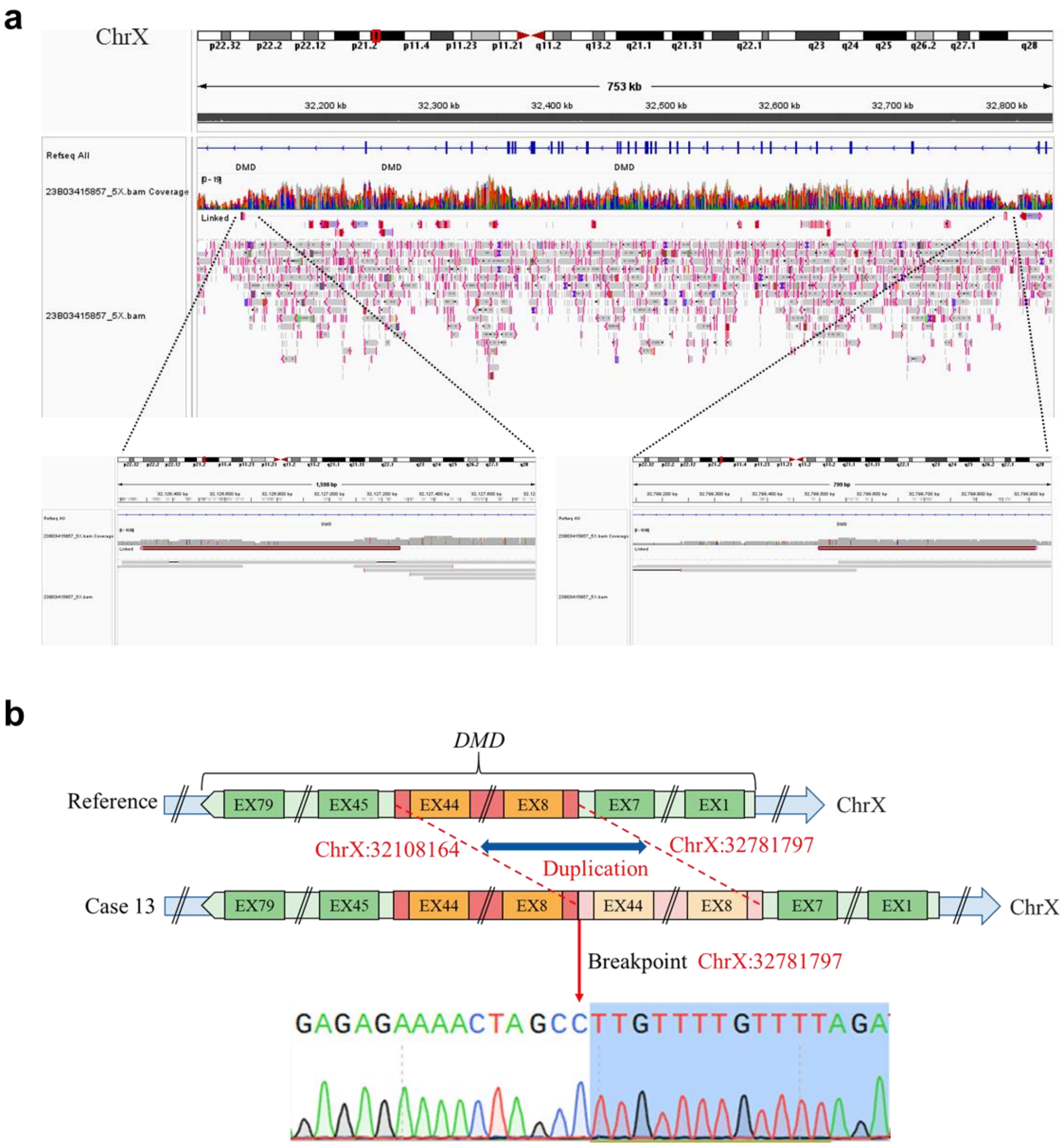


**Supplementary Figure 14**. Detection and confirmation of *DMD* gene duplication in case 13. **A.** Long-read genome sequencing data for case 13 was visualized using the IGV software. Reads supporting the duplication are shown in red. **B.** A tandem duplication of exons 8-44 (ChrX:32108164-32781797, hg38) in the *DMD* gene was identified in case 13 by Long-read genome sequencing and Sanger sequencing. The junction of breakpoint was located at chrX:32781797 (hg38). Chr, chromosome; EX, exon.

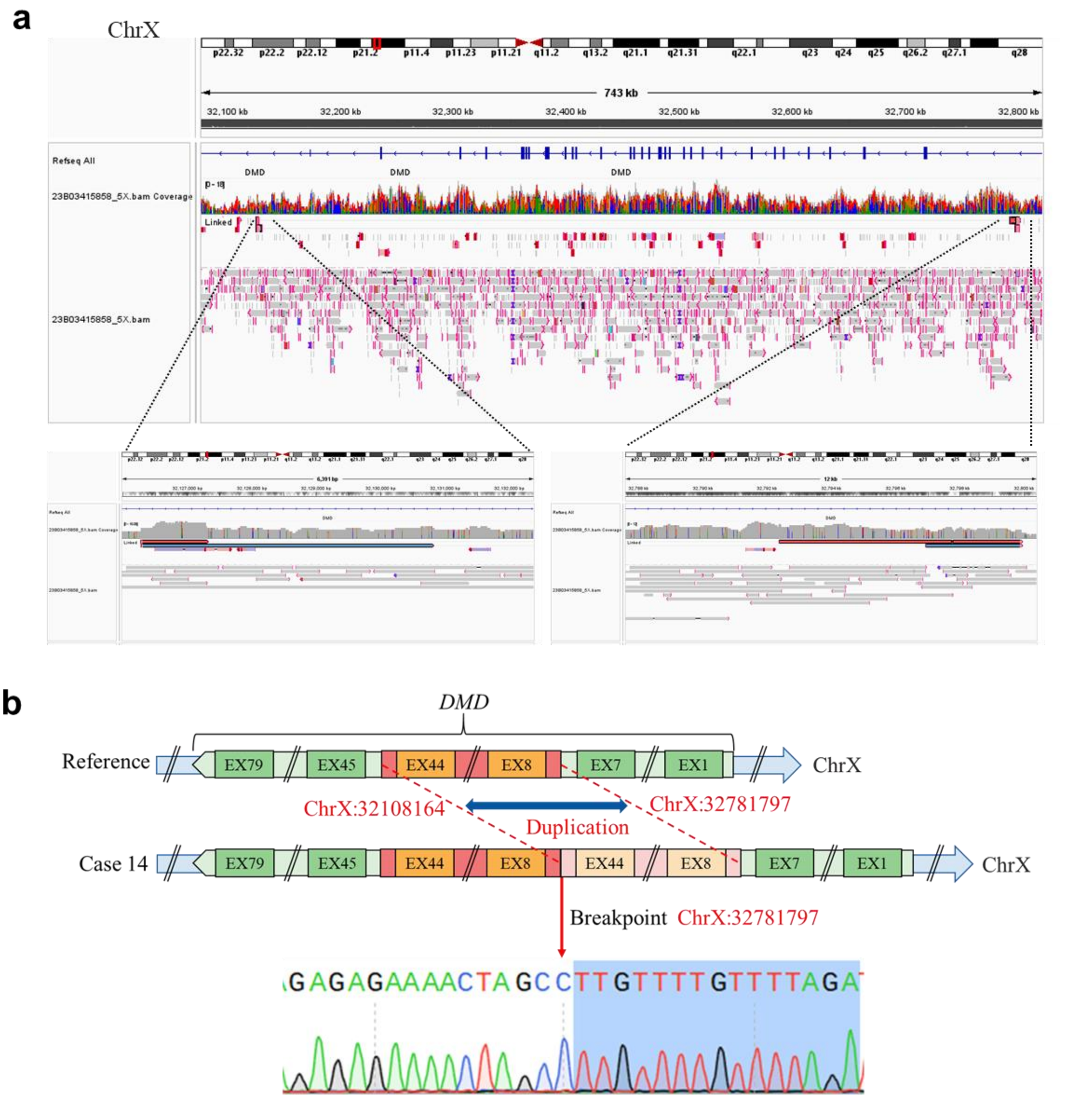


**Supplementary Figure 15**. Detection and confirmation of *DMD* gene duplication in case 14. **A.** Long-read genome sequencing data for case 14 was visualized using the IGV software. Reads supporting the duplication are shown in red or blue. **B.** A tandem duplication of exons 8-44 (ChrX:32108164-32781797, hg38) in the *DMD* gene was identified in case 14 by Long-read genome sequencing and Sanger sequencing. The junction of breakpoint was located at chrX:32781797 (hg38). Chr, chromosome; EX, exon.

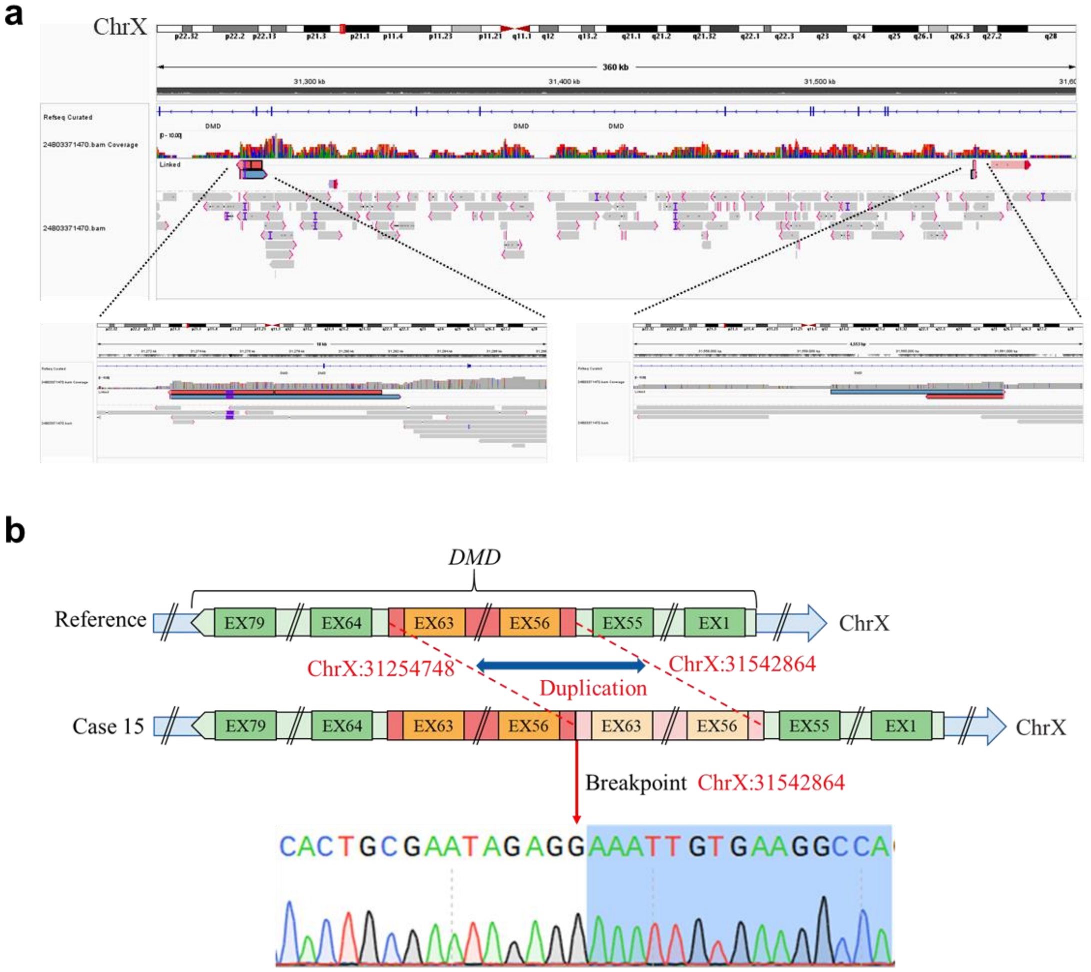


**Supplementary Figure 16**. Detection and confirmation of *DMD* gene duplication in case 15. **A.** Long-read genome sequencing data for case 15 was visualized using the IGV software. Reads supporting the duplication are shown in red or blue. **B.** A tandem duplication of exons 56-63 (ChrX:31254748-31542864, hg38) in the *DMD* gene was identified in case 15 by Long-read genome sequencing and Sanger sequencing. The junction of breakpoint was located at chrX:31542864 (hg38). Chr, chromosome; EX, exon.

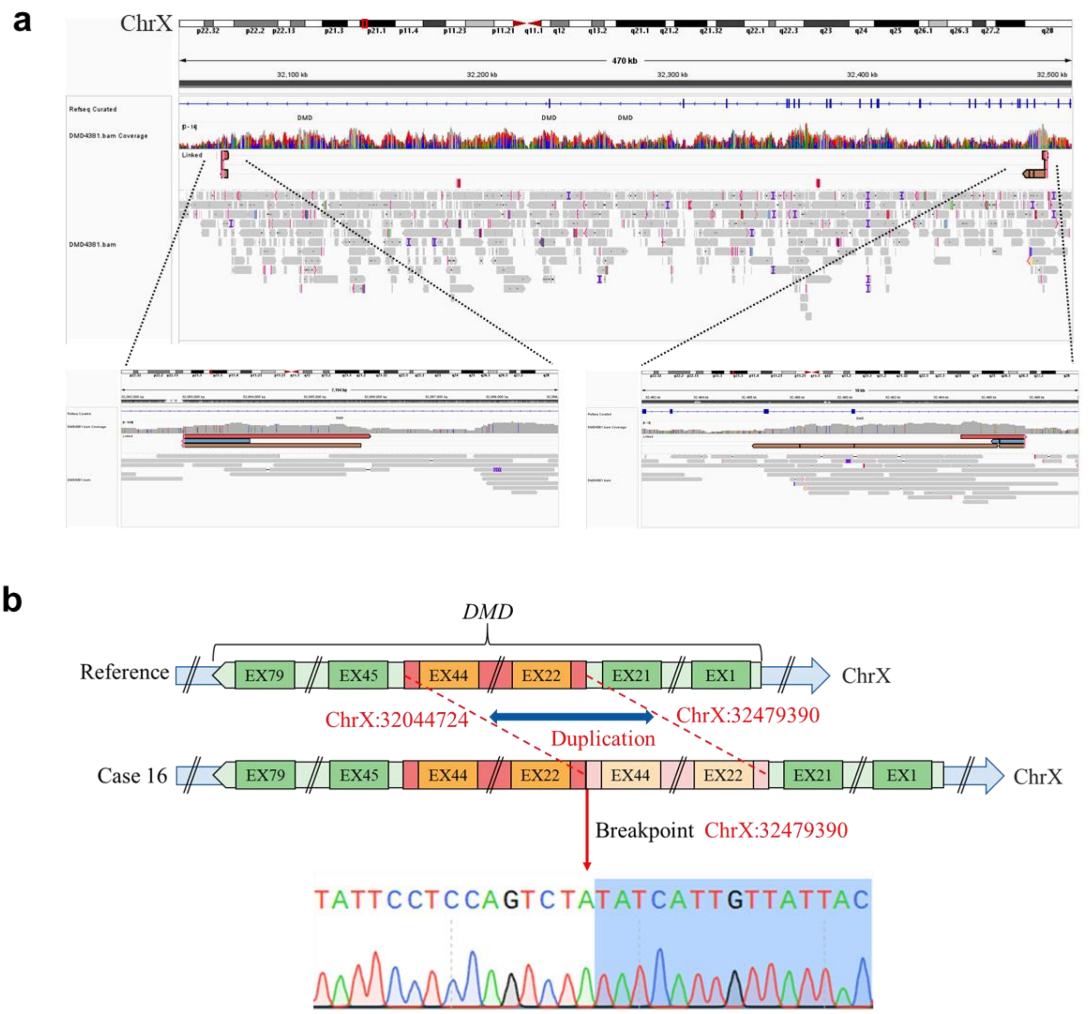


**Supplementary Figure 17**. Detection and confirmation of *DMD* gene duplication in case 16. **A.** Long-read genome sequencing data for case 16 was visualized using the IGV software. Reads supporting the duplication are shown in red, blue or brown. **B.** A tandem duplication of exons 22-44 (ChrX:32044724-32479390, hg38) in the *DMD* gene was identified in case 16 by Long-read genome sequencing and Sanger sequencing. The junction of breakpoint was located at chrX:32479390 (hg38). Chr, chromosome; EX, exon.

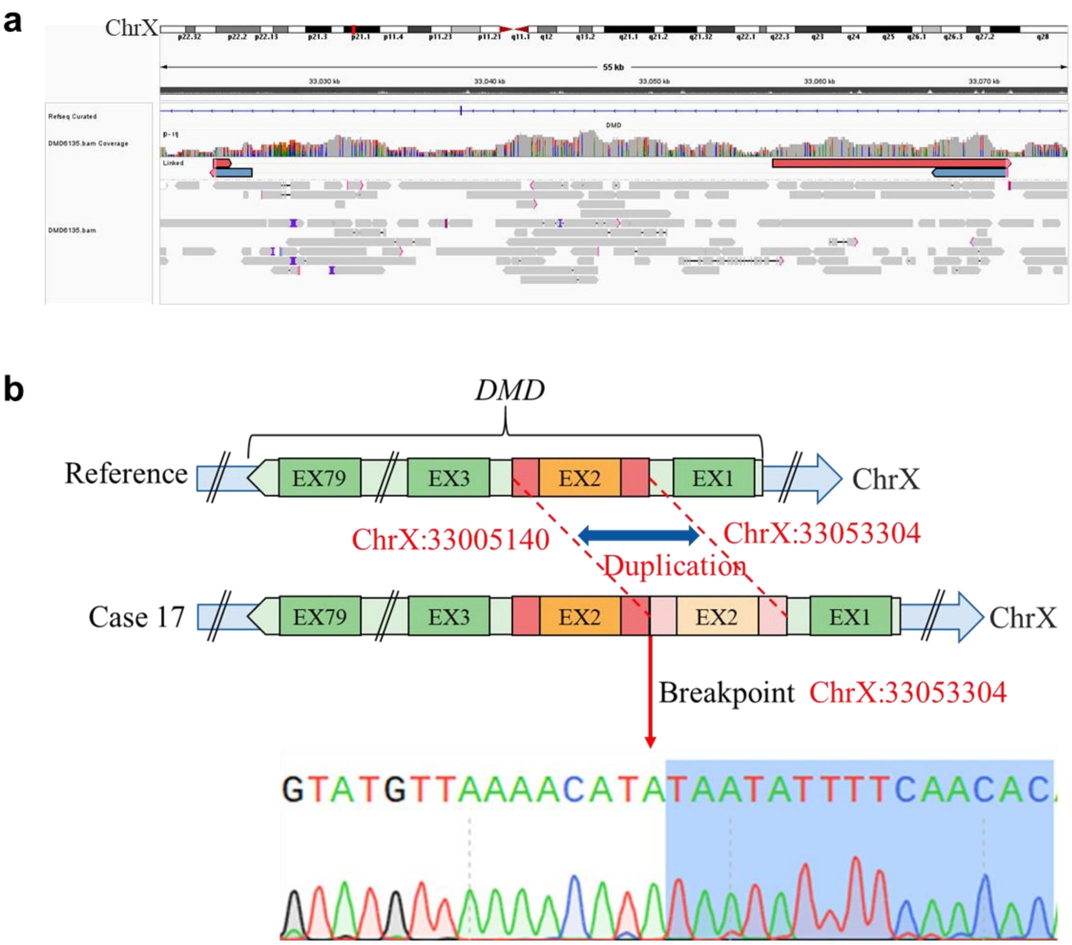


**Supplementary Figure 18**. Detection and confirmation of *DMD* gene duplication in case 17. **A.** Long-read genome sequencing data for case 17 was visualized using the IGV software. Reads supporting the duplication are shown in red or blue. **B.** A tandem duplication of exons 2 (ChrX:33005140-33053304, hg38) in the *DMD* gene was identified in case 17 by Long-read genome sequencing and Sanger sequencing. The junction of breakpoint was located at chrX:33053304 (hg38). Chr, chromosome; EX, exon.

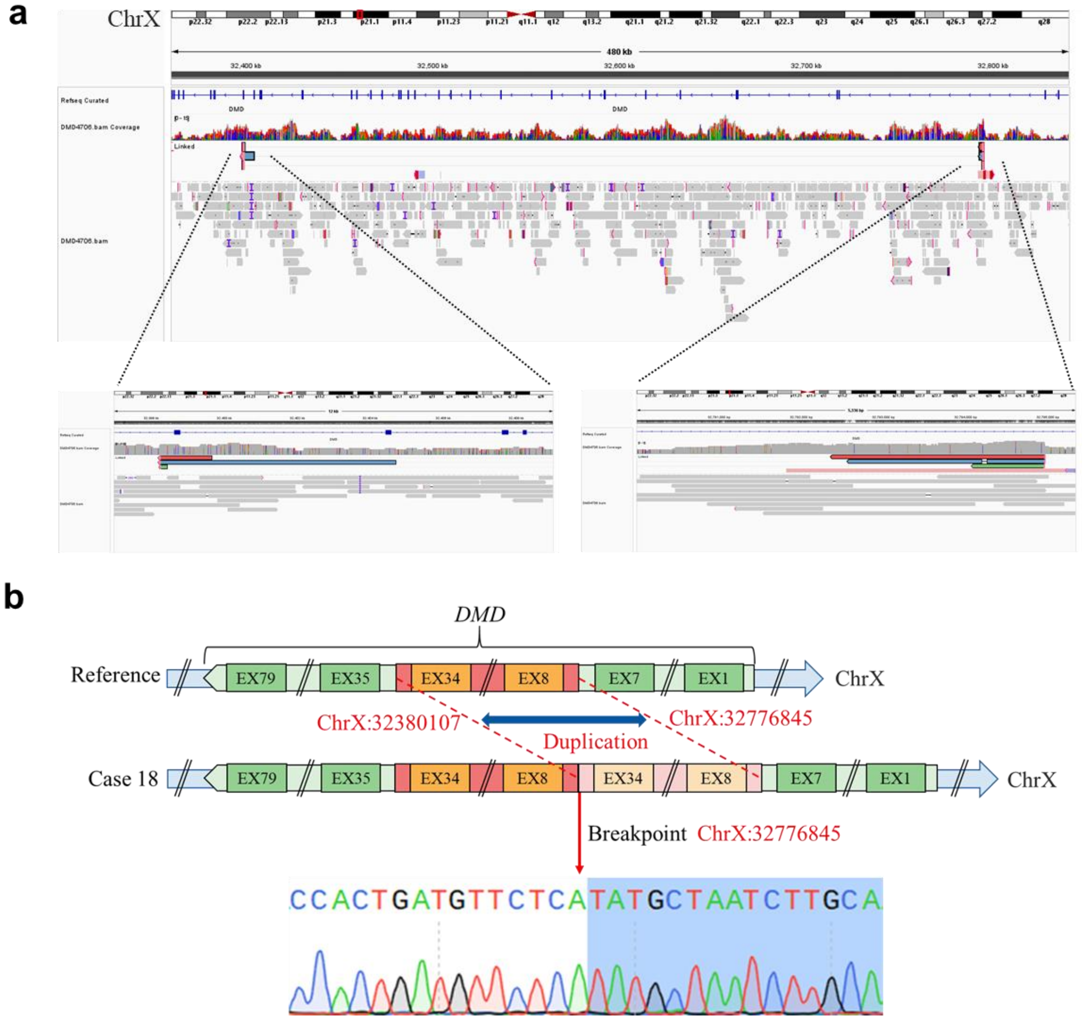


**Supplementary Figure 19**. Detection and confirmation of *DMD* gene duplication in case 16. **A.** Long-read genome sequencing data for case 16 was visualized using the IGV software. Reads supporting the duplication are shown in red, blue or green. **B.** A tandem duplication of exons 8-34 (ChrX:32380107-32776845, hg38) in the *DMD* gene was identified in case 16 by Long-read genome sequencing and Sanger sequencing. The junction of breakpoint was located at chrX:32776845 (hg38). Chr, chromosome; EX, exon.

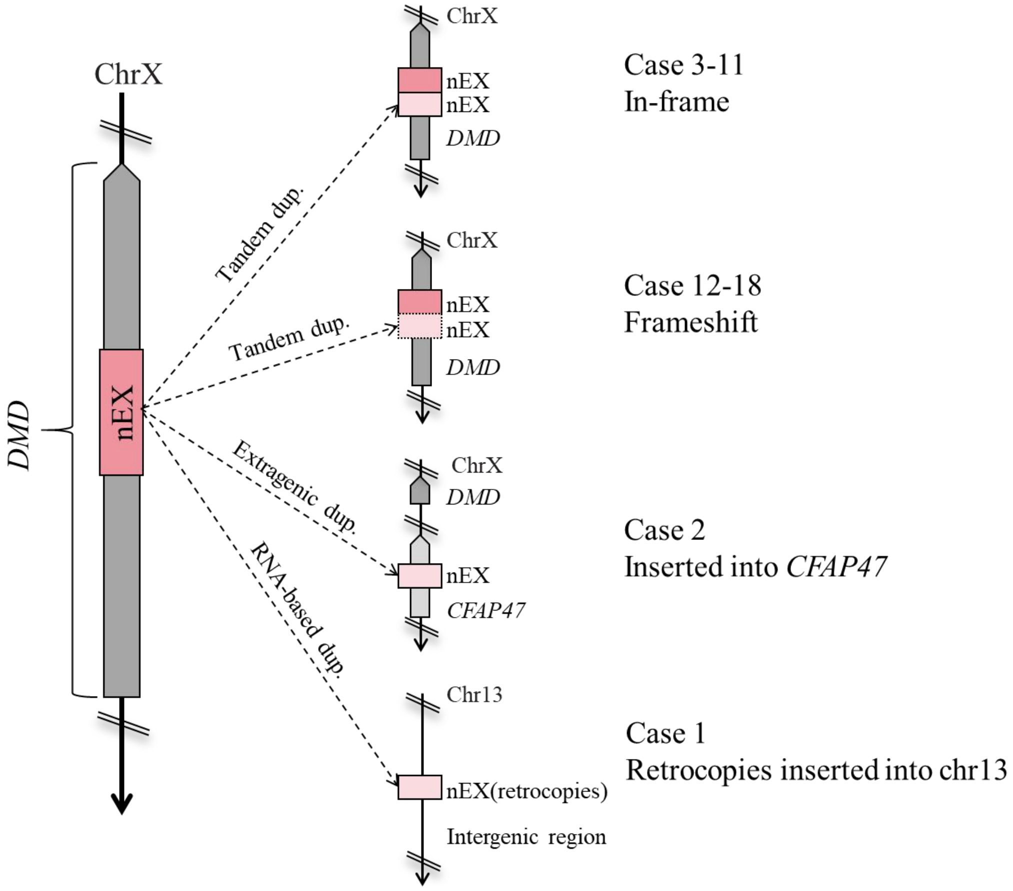


**Supplementary Figure 20**. Schematic representations of candidate structural variants resolved using long-read genome sequencing. In Case 3-11, the variation type is in-frame tandem duplication. In Case 12-18, the variation type is frameshift tandem duplication. In Case 2, the duplication is inserted in reverse into the *CFAP47* gene region without damaging the original *DMD* gene. In Case 1, a partial mRNA sequence of *DMD* is reverse transcribed to DNA and inserted into the intergene region of chr13 without damaging the original *DMD* gene. nEX, A fragment of the *DMD* gene encompassing n exons; nEX(retrocopies), Retrocopies containing n exons of the *DMD* gene; n, 1-79; EX, exon; chr., chromosome; Dup., duplication.

**Supplementary Table 1.** Eighteen carriers of *DMD* gene duplications were initially identified by short-read sequencing and confirmed by multiplex ligation-dependent probe amplification (MLPA) or qPCR.

| Case ID | Short read sequencing | MLPA | qPCR |
|---|---|---|---|
| 1 | EX30-37; 39-40 Dup. | EX29-31;32-37 | EX29;30;32-37;39;40, ratio=1.5 |
| 2 | EX56-EX60 Dup. | / | EX56;60, ratio=1.5 |
| 3 | EX2-19 Dup. | / | EX2;3;7;8;10;11;18;19, ratio=1.5 |
| 4 | EX34-36 Dup. | / | EX34;35;36, ratio=1.5 |
| 5 | EX10-23 Dup. | / | EX10;16;17;23, ratio=1.5 |
| 6 | EX48-79 Dup. | / | EX48;54;67;79, ratio=1.5 |
| 7 | EX54-60 Dup. | EX54-60, ratio=1.5 | / |
| 8 | EX54-60 Dup. | / | EX54;55;58;60, ratio=1.5 |
| 9 | EX6-16 Dup. | / | EX6;11;16, ratio=1.5 |
| 10 | EX75-79 Dup. | EX76-79, ratio=1.5 | / |
| 11 | EX37-44 Dup. | / | EX37;44, ratio=1.5 |
| 12 | EX22-44 Dup. | EX22-44, ratio=1.5 | / |
| 13 | EX8-44 Dup. | EX8-44, ratio=1.5 | / |
| 14 | EX8-44 Dup. | EX8-44, ratio=1.5 | / |
| 15 | EX56-63 Dup. | EX56-63, ratio=1.5 | / |
| 16 | EX22-44 Dup. | / | EX22;38;44, ratio=1.5 |
| 17 | EX2 Dup. | / | EX2, ratio=1.5 |
| 18 | EX8-34 Dup. | EX8-34， ratio=1.5 | / |

Dup., duplication; EX, exon; /, not testing.

**Supplementary Table 2**. Primers used for breakpoints validation

| Case ID | Prime F (5'-3') | Prime R (5'-3') |
|---|---|---|
| 1 | ccatcaaagttccacaagttcaaag | gtgaccaagcagcaaacttgat |
| 2 | caaaatgctgttcttctgagactggttg | ggtaggctgtcctaagaaaggaggatgac |
| 3 | catcagaatcaactgacgagct | agcacaaagctagccaagtt |
| 4 | gtgagactagctcatgctggt | ccttgatgtatagtcagagatgtcatct |
| 5 | atccatgagcatagaatgttcttcca | taagtaatgtgtcccttgagtggtc |
| 6 | tagttatctgttgtaactcagggtgacta | tgctcctggatgtgctgtt |
| 7 | tgtcagaccagcaaggacat | gacatcctacgcacatcgttg |
| 8 | tgtcagaccagcaaggacat | gacatcctacgcacatcgttg |
| 9 | agtcagttactcagtgaatacaggtc | cttcctgaaagctctccacgact |
| 10 | agtaggtagattggttcaggtcct | cctcacaatcaaggctaatgcagtt |
| 11 | cctttcagagtactgcgcaaccttcg | tcagcacccaaccgctcaagatacag |
| 12 | ttcacctcctggacaactgatt | actcactcactacactgaggac |
| 13 | cttgtgaatgtgtcagcctctctcctgc | gagaagaggttggggtagagagtttcagac |
| 14 | cttgtgaatgtgtcagcctctctcctgc | gagaagaggttggggtagagagtttcagac |
| 15 | catgcattcaccagagcagtttgacct | gtggttaaaagcctgtccttttctcacaat |
| 16 | ttcacctcctggacaactgatt | actcactcactacactgaggac |
| 17 | tacaaagacgcatgtccctaac | ctcttctgcttgcattgctcat |
| 18 | ccatcagatcacatgagcattcacacact | tgacagaatggctggcagct |

**Supplementary Table 3.** The inconsistency of the MLPA, qPCR, and short-read sequencing test results in Case 1

| *DMD* | MLPA | qPCR | NGS |
|---|---|---|---|
| EX29 | Dup. | Dup. | Negative |
| EX30 | Dup. | Dup. | Dup. |
| EX31 | Dup. | Negative | Dup. |
| EX32 | Negative | Dup. | Dup. |
| EX33 | Dup. | Dup. | Dup. |
| EX34 | Dup. | Dup. | Dup. |
| EX35 | Dup. | / | Dup. |
| EX36 | Dup. | / | Dup. |
| EX37 | Dup. | Dup. | Dup. |
| EX38 | Negative | Negative | Negative |
| EX39 | Negative | Dup. | Dup. |
| EX40 | Negative | Dup. | Dup. |

Dup., duplication; EX, exon; /, not testing.
